# Clinical Trials in COVID-19 Management & Prevention: A Meta-epidemiological Study examining methodological quality

**DOI:** 10.1101/2020.11.29.20237875

**Authors:** Kimia Honarmand, Jeremy Penn, Arnav Agarwal, Reed Siemieniuk, Romina Brignardello-Petersen, Jessica J Bartoszko, Dena Zeraatkar, Thomas Agoritsas, Karen Burns, Shannon M. Fernando, Farid Foroutan, Long Ge, Francois Lamontagne, Mario A Jimenez-Mora, Srinivas Murthy, Juan Jose Yepes Nuñez, Per O Vandvik, Zhikang Ye, Bram Rochwerg

## Abstract

**Background:** The coronavirus disease (Covid-19) pandemic has produced a large number of clinical trial reports with unprecedented rapidity, raising concerns about methodological quality and potential for research waste.

**Objectives:** To describe the characteristics of randomized clinical trials (RCTs) investigating prophylaxis or treatment of Covid-19 infection and examine the effect of trial characteristics on whether the study reported a statistically significant effect on the primary outcome(s).

**Study Design:** Meta-epidemiological study of Covid-19 treatment and prophylaxis RCTs.

**Eligibility criteria:** English-language RCTs (peer-reviewed or preprint) that evaluated pharmacologic agents or blood products compared to standard care, placebo, or an active comparator among participants with suspected or confirmed Covid-19 or at risk for Covid-19. We excluded trials of vaccines or traditional herbal medicines.

**Information sources:** We searched 25 databases in the US Centre for Disease Control Downloadable Database from January 1 to October 21, 2020.

**Trial appraisal and synthesis methods:** We extracted trial characteristics including number of centres, funding sources (industry versus non-industry), and sample size. We assessed risk of bias (RoB) using the modified Cochrane RoB 2.0 Tool. We used descriptive statistics to summarize trial characteristics and logistic regression to evaluate the association between RoB due to the randomization process, centre status (single vs. multicentre), funding source, and sample size, and statistically significant effect in the primary outcome.

**Results:** We included 91 RCTs (46,802 participants) evaluating Covid-19 therapeutic drugs (n = 76), blood products (n = 9) or prophylactic drugs (n = 6). Of these, 40 (44%) were single-centre, 23 (25.3%) enrolled < 50 patients, and 28 (30.8%) received industry funding. RoB varied across trials, with high or probably high overall RoB in 75 (82.4%) trials, most frequently due to deviations from the intended protocol (including blinding) and randomization processes. Thirty-eight trials (41.8%) found a statistically significant effect in the primary outcome. RoB due randomization (odds ratio [OR] 3.77, 95% confidence interval [CI], 1.47 to 9.72) and single centre trials (OR 3.15, 95% CI, 1.25 to 7.97) were associated with higher likelihood of finding a statistically significant effect.

**Conclusions:** There was high variability in RoB amongst Covid-19 trials. RoB attributed to the randomization process and single centre status were associated with a three-fold increase in the odds of finding a statistically significant effect. Researchers, funders, and knowledge users should remain cognizant of the impact of study characteristics, including RoB, on trial results when designing, conducting, and appraising Covid-19 trials.

## INTRODUCTION

The rapid rise in the number of cases, hospitalizations, and deaths due to Coronavirus disease 2019 (Covid-19) has been paralleled by an exponential rise in scientific publications related to Covid-19. The number publications with the terms ‘COVID-19’ or ‘SARS-CoV-2’ in their title or abstract was over 17,000 as of May 31^st^ and over 57,000 as of October 5^th^, 2020.

The global search to identify effective interventions against Covid-19 has led to an unprecedented rise in clinical trial activity worldwide. As of October 5, 2020, the World Health Organization (WHO) Global Coronavirus COVID-19 Clinical Trial Tracker reports that there are currently over 2,300 clinical trials at various stages of completion. The rapidity with which clinical trials in Covid-19 are being planned, completed, and disseminated has triggered concerns about their methodological quality.^1 2^ Flaws in study design may lead to biased estimates of intervention effects, leading to treatment decisions that are at best ineffectual, and at worst harmful to patients. The well-known waste in biomedical research may be enhanced by the COVID-19 pandemic.^3^

Several recent reports have described the design characteristics of registered trials of Covid-19 therapies.^3-8^ These reports, however, are based on registered trials, many of which will not proceed to completion and will therefore not impact clinical knowledge or practice. In addition, the appraisal of trial quality from registries does not include assessment of trial conduct as well as analysis.

We conducted a meta-epidemiological study of published Covid-19 randomized controlled trials (RCTs) to (1) describe trial characteristics, including risk of bias (RoB), and (2) evaluate the association between trial characteristics and the likelihood of finding statistically significant results for the primary outcome.

## METHODS

### Study Design

We performed this meta-epidemiological study as part of a living systematic review and network meta-analysis of RCTs examining Covid-19 prevention and therapy.^9^ We prepared this manuscript in accordance with the Preferred Reporting Items for Systematic Review and Meta-Analysis (PRISMA) statement.^10^

### Protocol Registration

We registered the protocol for this study in the Prospective Register of Systematic Reviews (PROSPERO 2020: CRD42020192095).

### Eligibility Criteria

We included English language RCTs of any publications status (peer-reviewed publication or preprint) that enrolled patients with suspected, probable or confirmed COVID-19, or at risk for contracting COVID-19, and compared the effect of pharmacologic agents or blood products against standard care, a placebo, or an active comparator (i.e., another pharmacologic agent or blood product). We excluded trials of vaccines or traditional herbal medicines that included more than one molecule or did not have a specific molecular weight dosing.

### Information Sources & Search Strategy

The complete search strategy is shown in **Supplementary Material Online 1**. We used the ongoing literature search performed by Centre for Disease Control (CDC), which includes 25 databases of published studies and repositories of unpublished studies (medRxiv and bioRxiv), to find potentially relevant articles of therapies related to SARS-CoV-2 and COVID-19 from January 1 to October 21, 2020. For pragmatic reasons, we excluded trials published in languages other than English.

### Study Selection

Working in pairs, reviewers screened, independently and in duplicate, titles and abstracts and then full-texts for articles found potentially eligible at the title and abstract screening stage. We resolved discrepancies by discussion and where needed, by third party adjudication.

### Data Extraction

Using a pre-developed data extraction form, we extracted study characteristics including: registration status (registered vs. non-registered), publication status (preprint vs. peer reviewed publication), trial design (single-centre or multicentre), funding source (industry vs. non-industry), study interventions (number of study arms, intervention details, type of comparator [active vs. not]). We also extracted details about the trial’s reported primary outcome(s), including whether the outcome was binary vs. continuous vs. ordinal, patient-important or surrogate, event rates and summary statistics for binary and continuous outcomes, respectively, and whether there was a statistically significant difference detected in the primary outcome. For trials that reported more than one primary outcome, we recorded the primary outcome with the highest effect size.

### Risk of Bias Assessment

Three reviewers evaluated RoB of included studies using the modified version of the Cochrane RoB 2.0 tool independently (**Supplementary Material Online 2**). Discrepancies were resolved by consensus. The modified Cochrane RoB tool rates methodological quality of each included study as low, probably low, probably high, or high RoB across each of five domains, reflecting bias: (1) from the randomization process, (2) due to deviations from the intended intervention (which included blinding procedures), (3) due to missing data, (4) due to measurement of the outcome, and (5) in selection of the reported results. We categorized overall study RoB as the highest rating across any of the five domains.

### Data Analysis

We used descriptive statistics (means and standard deviations, medians and interquartile ranges, and proportions and confidence intervals, as appropriate) to summarize trial characteristics and RoB for the included trials.

We then conducted logistic regression analyses to assess the association between a trial finding a statistically significant effect (defined as a p-value equal to or less than 0.05) and pre-specified trial characteristics, including:

- RoB due to randomization: dichotomized into low/ probably low RoB and high/ probably high RoB
- Centre status: Multicentre vs. single centre trial
- Funding source: those with any industry funding vs. those without industry funding
- Trial sample size (using the total number randomized as a continuous variable)

We selected these trial characteristics *a priori* based on the hypothesis that these specific trial characteristics were most important in influencing trial findings. We included RoB due to the randomization process, as opposed to other RoB domains, as we anticipated the randomization process to have the highest association with trial outcomes and due to the anticipated limited variability between trials in other RoB domains, which would not allow for meaningful interpretation or conclusions.

Among the four selected predictor variables, we used purposeful selection of predictor variables according to the approach described by **Bursac and colleagues**.^11^ The process began with univariate analysis of each of the four pre-specified predictors. Then, variables that yield a p-value of less than 0.25 are selected as candidates for the multivariable analysis and entered into the model. Through an iterative process of variable selection, variables are retained in the model only if they (1) have an association with the outcome as defined by a p-value of < 0.1 or (2) have a confounding effect, defined a change in the group coefficient by more than 15% when the variable is removed as compared to the full model. This approach allows for iterative selection of predictor variables and retains in the model those predictors that are not themselves significantly associated with the outcome but contribute to the effect of other predictors. We planned to perform subgroup analyses to evaluate the impact of trial characteristics on trial outcomes among trials that were preprints compared to those published in peer-reviewed journals but the relatively small number of trials prohibited this analysis. We used Statistical Package for the Social Sciences (SPSS) version 26.0 (IBM Corporation) for all descriptive and regression analyses and Stata/IC 16.1 (StataCorp LLC) to produce the forest plot of effect sizes.

## RESULTS

### Study Selection

The search identified 13,536 records which were reviewed in duplicate as part of a living network meta-analysis,^9^ and yielded 103 trials of therapeutic or prophylactic interventions for Covid-19. We excluded five RCTs published in languages other than English, two trials that reported on a cohort overlapping with another included trial, two that reported preliminary results but not findings related to their primary outcomes, and three unpublished studies that were included in a meta-analysis with insufficient information to include in our review. We included a total of 91 clinical trials (54 peer-reviewed publications, 37 preprints) in this analysis.

### Trial Characteristics

#### Overall trial characteristics

**Table 1** presents the aggregate characteristics of included studies. The 91 included trials enrolled a total of 46,802 patients between January 18 (first recruitment) and October 4 (last recruitment). Included trials evaluated one or more drugs (n = 76,^12-87^)or blood products (n = 9,^88-96^) to treat patients with suspected or confirmed Covid-19 or drugs used as prophylaxis for patients at risk for Covid-19 (n = 6,^97-102^). All but one of the trials were parallel group design (one trial was a cluster randomized design). Thirty of 91 trials were conducted by a country in the Western Pacific Region, primarily China (n = 27). **Figure 1** illustrates the proportion of trials that were led by countries in various regions, as defined by the WHO. All but three trials were pre-registered. Fifty-one trials were multicentre whereas 40 were single centre. Trial sample size ranged from 10 to 14,247 (median: 84, interquartile range [IQR]: 151); 23 trials enrolled less than 50 patients, 51 enrolled 50 to 400 patients, and 17 enrolled over 400 patients.

**Figure 1:**
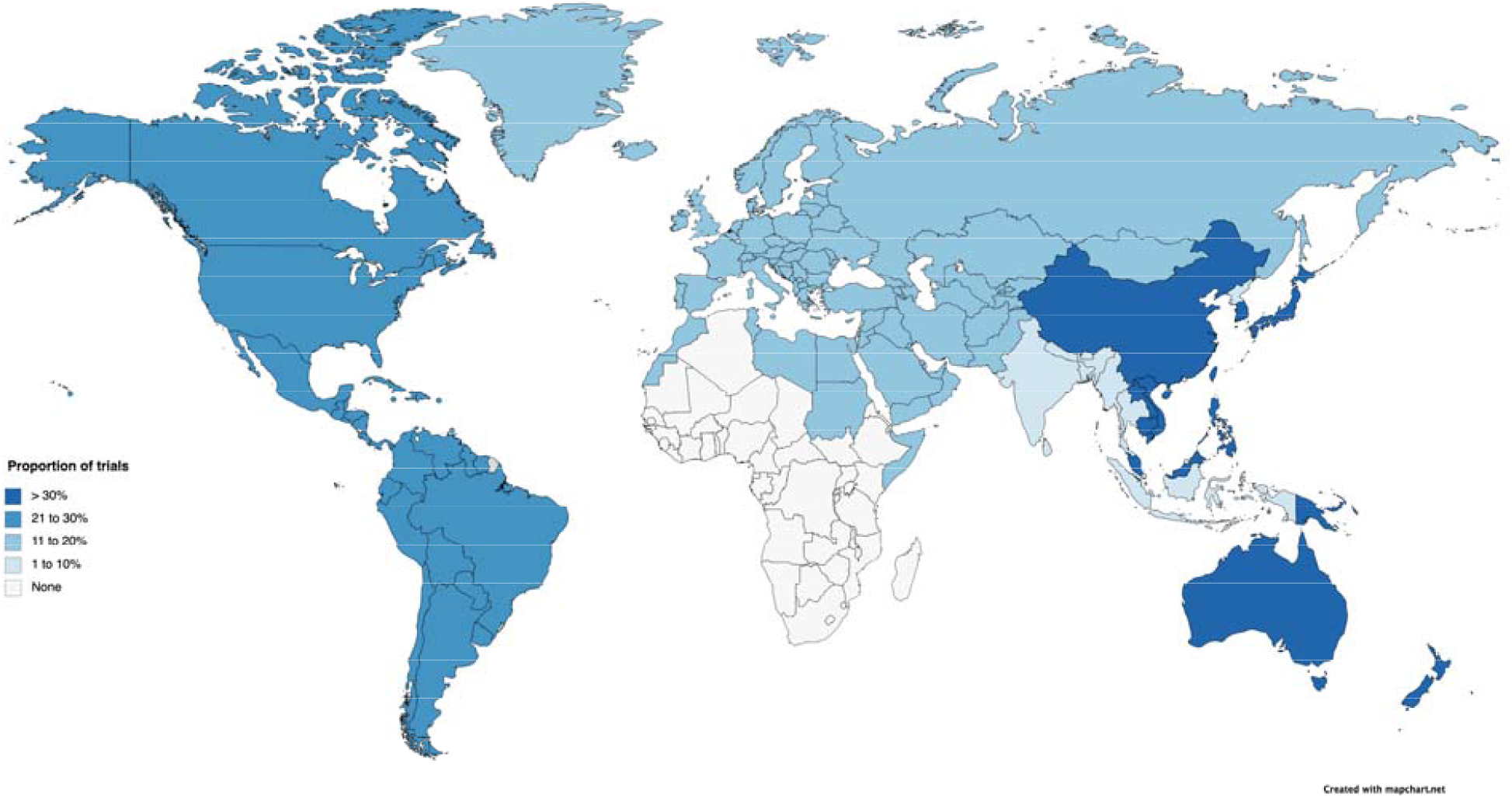
The geographical distribution of trials according to WHO region.

**Table 1.**
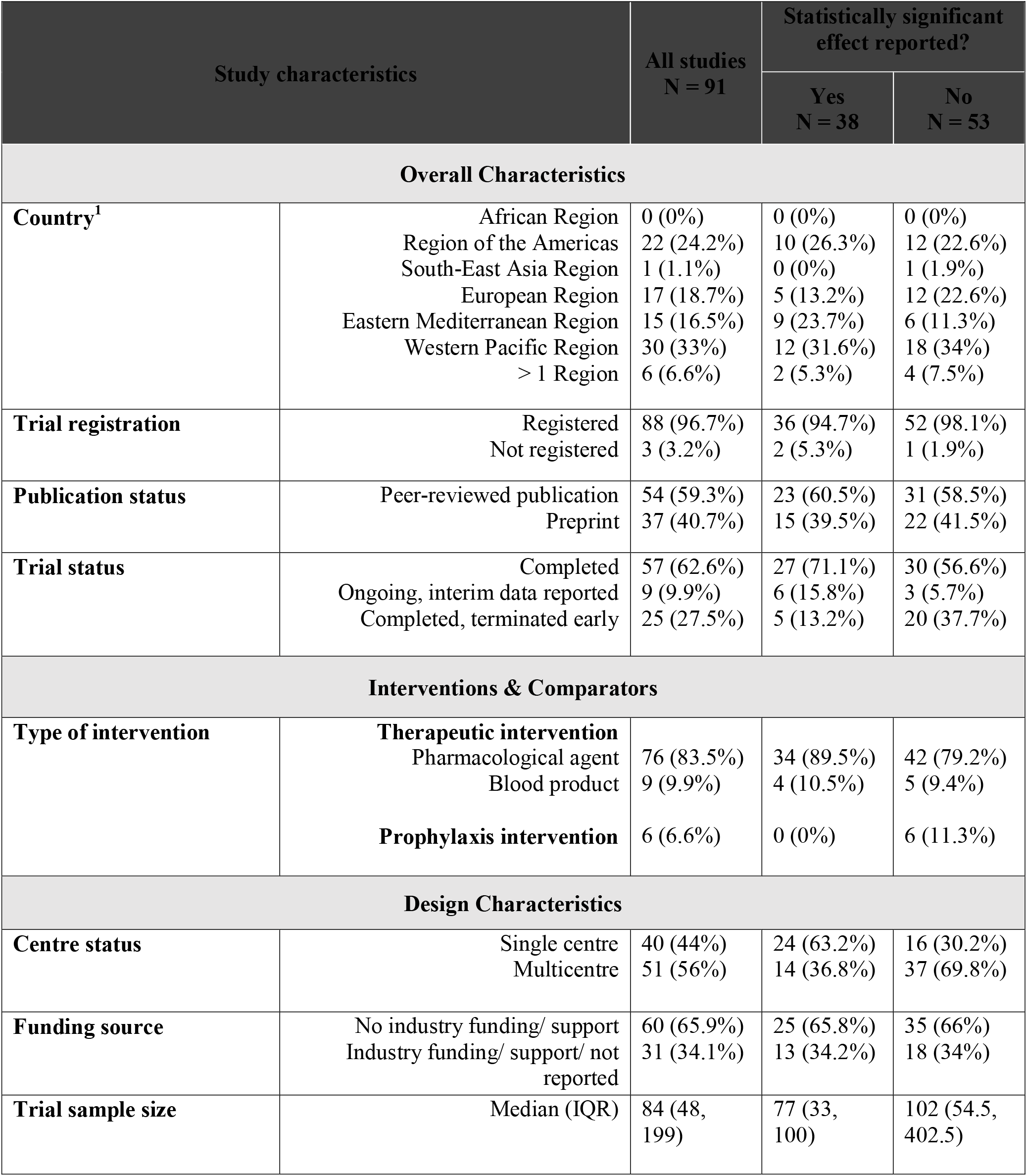
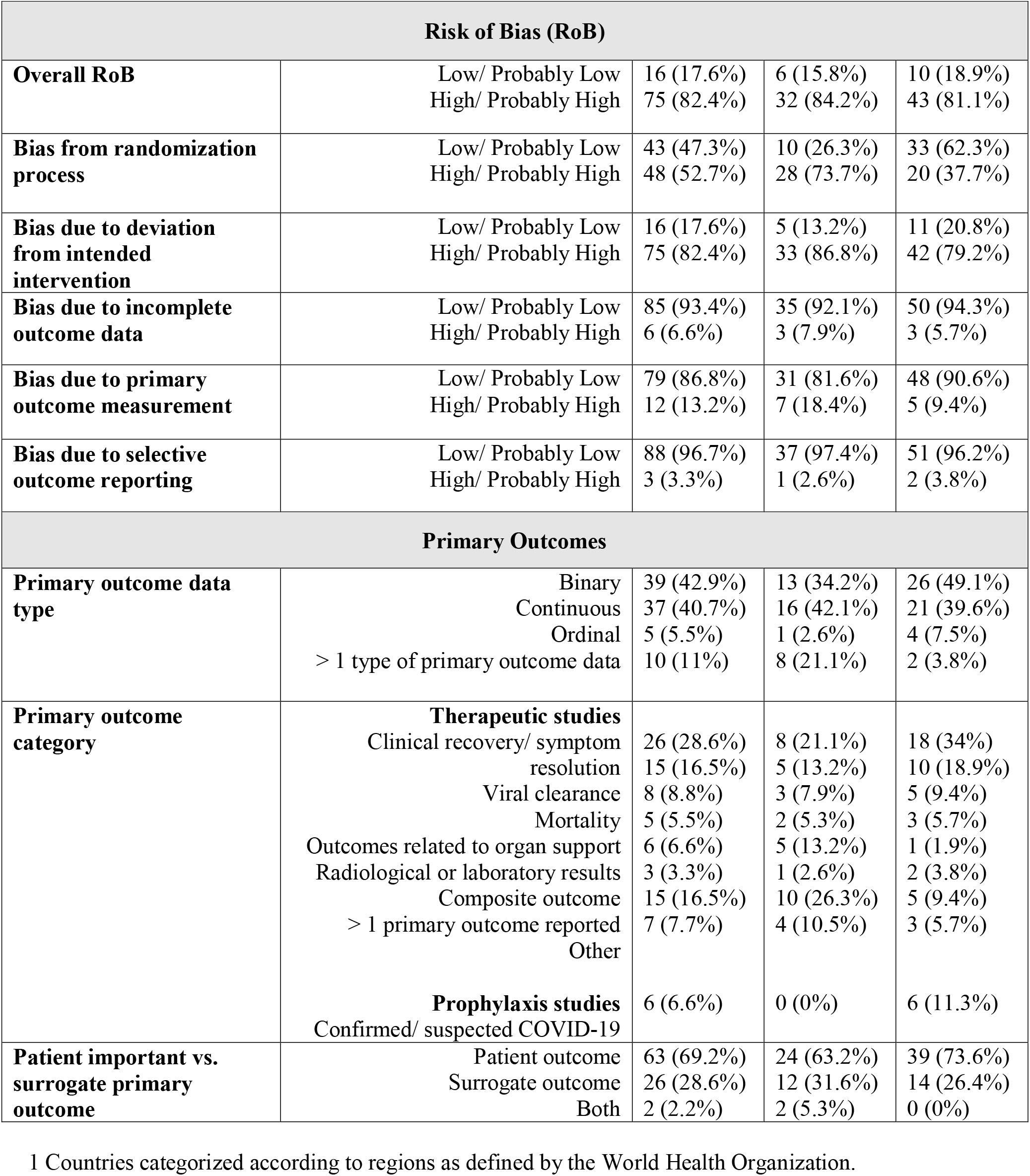
Study characteristics & risk of bias.

Among 88 studies that reported their funding source, 28 received at least some industry support including complete industry funding in 10 trials, partial industry funding for 7 trials, and provision of intervention/ medications by industry in 11. The 60 trials that reported no industry support were funded by governmental sources (n = 31), academic institutions (n = 9), multiple sources (government, academic institutional, and/ or not-for-profit organization; n = 13) or received no funding (n = 7).

#### Trial risk of bias

There was variability across various RoB domains. Seventy-five (82.4%) having overall high or probably high RoB (**Table 1**). Across individual RoB domains, there was high/ probably high RoB from the randomization process in 48 trials (52.7%), due to deviations from the intended protocol (which incorporates blinding procedures) in 75 (82.4%), due to incomplete primary outcome data in 6 (6.6%), due to incomplete primary outcome measurement in 12 (13.2%), and due to selective outcome reporting in 3 (3.3%; **Table 1**).

#### Trial primary outcomes

Table 1 presents the primary outcomes of included studies and their characteristics. The primary outcomes were binary in 39 trials, continuous in 37, ordinal in 5, and the remaining 10 trials reported more than one primary outcome. Among the 85 therapy trials, most trials (26 or 28.6%) reported a measure of clinical recovery or symptom resolution as the primary outcome. Thirty-eight studies reported a statistically significant effect (41.8%) and 53 reported no statistically significant difference (58.2%; **Table 1**).

### Association between trial characteristics and findings

We evaluated the association between each of the pre-specified trial characteristics on trial findings (whether or not a statistically significant effect was found). Bias due to the randomization process was high or probably high in 28 of 38 (73.7%) of trials that found a statistically significant effect on their primary outcome, compared with 20 of 53 (37.7%) of trials that found no statistically significant effect. **Figure 2** shows the RoB across the five domains on the modified Cochrane RoB tool across the two groups of trials.

**Figure 2:**
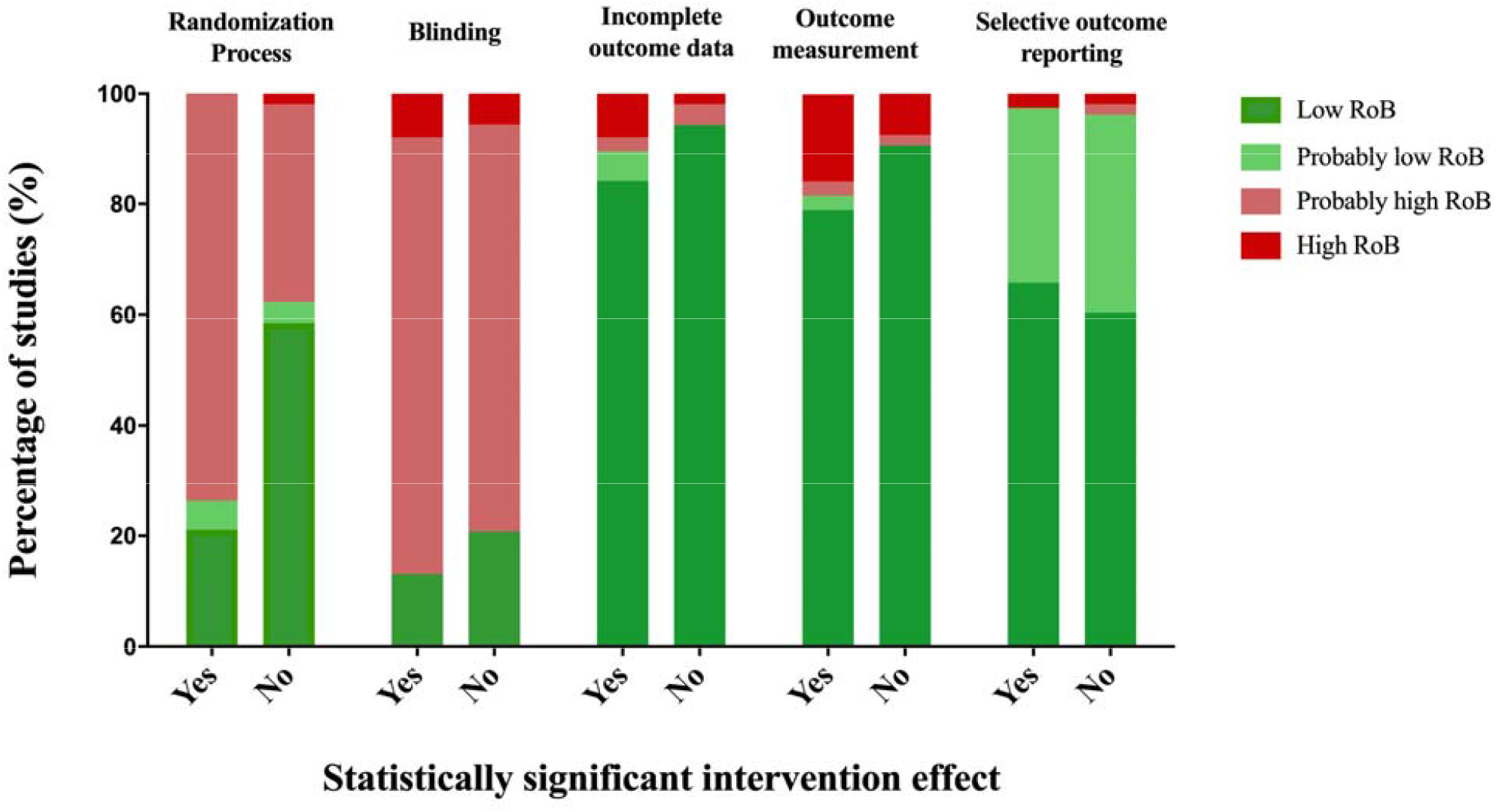
Risk of bias according to trial finding.

Single centre studies accounted for 24 of 38 (63.2%) trials that reported a statistically significant effect compared with 16 of 53 (30.2%) trials that reported no statistically significant effect (OR 3.93, 95% CI, 1.38 to 11.19). Thirteen of 38 trials (34.2%) that found a statistically significant effect were industry funded compared with 18 of 53 (34.0%) trials that found no statistically significant effect (OR 1.82, 95% CI, 0.61 to 5.43). Median sample size was 77 (IQR: 67) among trials that found a statistically significant effect and 102 (IQR: 348) in trials that found no statistically significant effect (OR 1.00 per patient randomized, 95% CI: 1.00 to 1.00, p = 0.74). Bias due to the randomization process was associated with higher odds of finding a statistically significant effect (OR 3.89, 95% CI, 1.46 to 10.36).

In univariate analysis, only bias due to the randomization process was associated with trial outcome (whether or not a statistically significant intervention effect was found); there was no association between trial outcome and centre status, funding source, and sample size (**Table 2**). In multivariable analysis, we found that higher bias due to the randomization process (OR 3.77, 95% CI, 1.47 to 9.72) and single centre trial status (OR 3.15, 95% CI, 1.25 to 7.97) were predictors of a trial finding a statistically significant effect.

**Table 2.** Association between trial characteristics and statistically significant results in primary outcome of Covid-19 clinical trials.

| Predictor variables | Univariable analysis |  | Multivariable model |  |
| --- | --- | --- | --- | --- |
|  | OR (95% CI) | p | OR (95% CI) | p |
| RoB due to randomization process<sup>1</sup> | 3.89 (1.46 to 10.36) | 0.01 | 3.77 (1.47 to 9.72) | 0.01 |
| Single centre vs. multicentre | 3.93 (1.38 to 11.19) | 0.01 | 3.15 (1.25 to 7.97) | 0.02 |
| Industry vs. non-industry support | 1.82 (0.61 to 5.43) | 0.28 | - | - |
| Total sample size | 1.00 (1.00 to 1.00) | 0.76 | - | - |

RoB: risk of bias; OR: odds ratio; CI: confidence interval
<sup>1</sup> Dichotomized into low/ probably low vs. high/ probably high

## DISCUSSION

In this meta-epidemiological study of clinical trials of Covid-19 prophylaxis and treatments, we found that 82.4% of trials had high or probably high RoB, 82.4% due to deviations from the intended intervention (including blinding) and 52.7% due to the randomization process (including allocation concealment and adequacy of the randomization procedure). Other trial characteristics were highly variable across studies: 44% were single centre trials, slightly less than one-third received at least some support from an industry source and all but 3 trials were registered in advance. Sample sizes were highly variable across studies, ranging from 10 to over 14,247, with one-quarter enrolling less than 50 patients.

The Covid-19 pandemic has seen the global research community embark on a collective search to identify effective prophylactic and therapeutic interventions against the disease. This global response has substantially exceeded that of previous pandemics: in the first six months, thousands of clinical trials had already been registered and hundreds were underway, compared with 71 registered trials after the onset of the H1N1/09 virus pandemic in 2009 and no registered trials after the Severe Acute Respiratory Syndrome (SARS) and Middle East Respiratory Syndrome (MERS) epidemics during the same time frame.^103^ This pandemic has also seen an unprecedented level of public interest. Early research findings are now routinely disseminated by researchers in preprint form (bypassing the long-held tradition of peer-review process), and on social media by mainstream media and the healthcare community. In most cases, this is done with inadequate attention to issues related to study design and methodologic quality.

Trial characteristics, including RoB, lead to low quality evidence, which may be uninformative at best and may cause harm to patients. In addition, poor quality trials absorb a disproportionate amount of attention from the general public and divert attention and research resources (i.e., efforts, financial support) away from other interventions which may be beneficial but remain under-investigated. These concerns are undoubtedly compounded when we consider the research resources allocated observational studies and RCTs that remain unpublished. The ultimate effect may be diminished public confidence in the scientific process, especially as data from low quality trials may not be reproducible and likely to be contradicted in subsequent, well-designed trials. In this study, we found that bias due to randomization process and single centre trial status were associated with increased odds of finding a statistically significant effect on the primary outcome, independent of the effect of sample size or industry funding source.

These findings highlight the need for researchers to do everything possible to minimize the risk of misleading trial results by prioritizing rigour in trial design (often competing with expediency), with particular focus on the randomization process. We also found that single centre trials were more likely to report a statistically significant interventional effect relative to multicentre trials, independent of the effect of sample size. The lack of an association between industry funding and the likelihood of finding a statistically significant effect is consistent with the findings of some previous meta-epidemiological studies,^104 105^ but inconsistent with other studies that found that industry funded trials are more likely to report a statistically significant effect.^106 107^ Lastly, we found no association between sample size and the likelihood of a statistically significant effect. While a previous meta-epidemiological study showed that small studies tend to overestimate effect sizes,^108^ that study also found that smaller trials had higher RoB across all domains, which may be the more likely explanatory variable.

This study has several strengths. We performed a comprehensive search as part of a living systematic review and NMA peer-reviewed and published in the BMJ, searched a large number of databases, included all Covid-19 RCTs examining drugs or blood products as therapeutics as well as drugs for prophylaxis. This living systematic review is currently informing the WHO living guidelines performed in collaboration with the MAGIC Evidence Ecosystem Foundation.^109^ The linkage to these trustworthy guidelines adds further rigor to the assessments of RoB through involvement of methodologists and unconflicted clinical experts making use of GRADE evidence summaries from the systematic review. In addition, we conducted RoB evaluation in duplicate, carefully assessed other trial characteristics that could influence likelihood of findings a statistically significant result. This study has several limitations. First, we did not include non-English trials which may influence the association between trial characteristics and trial outcomes. Furthermore, the relatively small sample of RCTs precluded our ability to conduct pre-planned subgroup analyses to evaluate the impact of trial characteristics on trial outcomes among trials that were preprints compared to those published in peer-reviewed journals. As such, updates on this report as more trials are published will allow for evaluation of a broader range of trial design characteristics and subgroup analyses to further understand the association between trial characteristics and trial outcomes.

## CONCLUSION

We found high variability in RoB amongst covid-19 trials across various RoB domains. RoB due to the randomization process and single centre status were associated with a three-fold increase in the odds of a trial finding a statistically significant effect. In their design and planning of Covid-19 trials, researchers are encouraged to consider the impact of trial characteristics and strive to generate reliable, high quality evidence. Funders should be cognizant of the ongoing research waste in Covid-19, limiting their support to well-designed trials that are likely to yield reliable, high quality evidence. Knowledge users should consider these findings when critically appraising and applying the findings of Covid-19 trials.

## Supporting information

PRISMA

## Data Availability

No additional data available.

## Ethical approval

Not applicable. All the work was developed using published/pre-print data.

## Contributors

KH and BR conceptualized the study. RS, RB, JJB, DZ, TA, KB, SMF, FF, LG, FL, MAJ, SM, JJY, PV, and ZY provided methodological guidance in study design, implementation, and interpretation. JJB led the systematic review search and study selection. DZ led data extraction and risk of bias evaluation. KH, JP, and AA collected additional data. FF and LG provided guidance on statistical analysis. KH performed data analysis. KH and BR drafted the manuscript. All authors approved the final version of the manuscript. KH is the guarantor. The corresponding author attests that all listed authors meet authorship criteria and that no others meeting the criteria have been omitted.

## Transparency declaration

KH affirms that this manuscript is an honest, accurate, and transparent account of the study being reported; that no important aspects of the study have been omitted; and that any discrepancies from the study as planned have been explained.

## Dissemination declaration

It is not applicable to disseminate the results to study participants and or patient organisations.

## Data sharing

No additional data available.

## Provenance and peer review

Not commissioned; externally peer reviewed.

## Patient and public involvement statement

It was not appropriate or possible to involve patients or the public in the design, or conduct, or reporting, or dissemination plans of our research.

The Corresponding Author has the right to grant on behalf of all authors and does grant on behalf of all authors, an exclusive licence (or non exclusive for government employees) on a worldwide basis to the BMJ Publishing Group Ltd to permit this article (if accepted) to be published in BMJ editions and any other BMJPGL products and sublicences such use and exploit all subsidiary rights, as set out in our licence.

This is an Open Access article distributed in accordance with the Creative Commons Attribution Non Commercial (CC BY-NC 4.0) license, which permits others to distribute, remix, adapt, build upon this work non-commercially, and license their derivative works on different terms, provided the original work is properly cited and the use is non-commercial. See: http://creativecommons.org/licenses/by-nc/4.0/.

