## Supplementary Material Online 1 for "Clinical Trials in COVID-19 Management & Prevention: A Meta-epidemiological Study examining methodological quality"

**Search Strategy:**

| Database | Strategy |
|---|---|
| Medline (Ovid) 1946- | (coronavir* OR corona virus* OR betacoronavir* OR covid19 OR covid 19 OR nCoV OR novel CoV OR CoV 2 OR CoV2 OR sarscov2 OR 2019nCoV OR wuhan virus*).mp. OR ((wuhan OR hubei OR huanan) AND (severe acute respiratory OR pneumonia*) AND outbreak*).mp. OR Coronavirus Infections/ OR Coronavirus/ OR betacoronavirus/<br><br>Limits: 2020-<br><br>OR<br><br>(novel coronavir* OR novel corona virus* OR covid19 OR covid 19 OR nCoV OR novel CoV OR CoV 2 OR CoV2 OR sarscov2 OR 2019nCoV OR wuhan virus*).mp. OR ((wuhan OR hubei OR huanan) AND (severe acute respiratory OR pneumonia*) AND outbreak*).mp. OR ((wuhan OR hubei OR huanan) AND (coronavir* OR betacoronavir*)).mp.<br><br>Limits: 2019- |
| Embase (Ovid) 1947- | (coronavir* OR corona virus* OR betacoronavir* OR covid19 OR covid 19 OR nCoV OR novel CoV OR CoV 2 OR CoV2 OR sarscov2 OR 2019nCoV OR wuhan virus*).mp. OR ((wuhan OR hubei OR huanan) AND (severe acute respiratory OR pneumonia*) AND outbreak*).mp. OR Coronavirus infection/ OR coronavirinae/ OR exp betacoronavirus/<br><br>Limits: 2020-<br><br>OR<br><br>(novel coronavir* OR novel corona virus* OR covid19 OR covid 19 OR nCoV OR novel CoV OR CoV 2 OR CoV2 OR sarscov2 OR 2019nCoV OR wuhan virus*).mp. OR ((wuhan OR hubei OR huanan) AND (severe acute respiratory OR pneumonia*) AND outbreak*).mp. OR ((wuhan OR hubei OR huanan) AND (coronavir* OR betacoronavir*)).mp.<br><br>Limits: 2019- |
| CAB Abstracts (Ovid) 1910- | (coronavir* OR corona virus* OR betacoronavir* OR covid19 OR covid 19 OR nCoV OR novel CoV OR CoV 2 OR CoV2 OR sarscov2 OR 2019nCoV OR wuhan virus*).mp. OR ((wuhan OR |

| | |
|---|---|
| | hubei OR huanan) AND (severe acute respiratory OR pneumonia*) AND outbreak*).mp. OR exp Betacoronavirus/<br><br>Limits: 2020-<br><br>OR<br><br>(novel coronavir* OR novel corona virus* OR covid19 OR covid 19 OR nCoV OR novel CoV OR CoV 2 OR CoV2 OR sarscov2 OR 2019nCoV OR wuhan virus*).mp. OR ((wuhan OR hubei OR huanan) AND (severe acute respiratory OR pneumonia*) AND outbreak*).mp. OR ((wuhan OR hubei OR huanan) AND (coronavir* OR betacoronavir*)).mp.<br><br>Limits: 2019- |
| Global Health (Ovid) 1910- | (coronavir* OR corona virus* OR betacoronavir* OR covid19 OR covid 19 OR nCoV OR novel CoV OR CoV 2 OR CoV2 OR sarscov2 OR 2019nCoV OR wuhan virus*).mp. OR ((wuhan OR hubei OR huanan) AND (severe acute respiratory OR pneumonia*) AND outbreak*).mp. OR exp Betacoronavirus/<br><br>Limits: 2020-<br><br>OR<br><br>(novel coronavir* OR novel corona virus* OR covid19 OR covid 19 OR nCoV OR novel CoV OR CoV 2 OR CoV2 OR sarscov2 OR 2019nCoV OR wuhan virus*).mp. OR ((wuhan OR hubei OR huanan) AND (severe acute respiratory OR pneumonia*) AND outbreak*).mp. OR ((wuhan OR hubei OR huanan) AND (coronavir* OR betacoronavir*)).mp.<br><br>Limits: 2019- |
| PsycInfo (Ovid) 1806- | (coronavir* OR corona virus* OR betacoronavir* OR covid19 OR covid 19 OR nCoV OR novel CoV OR CoV 2 OR CoV2 OR sarscov2 OR 2019nCoV OR wuhan virus*).mp. OR ((wuhan OR hubei OR huanan) AND (severe acute respiratory OR pneumonia*) AND outbreak*).mp.<br><br>Limits: 2020-<br><br>OR<br><br>(novel coronavir* OR novel corona virus* OR covid19 OR covid 19 OR nCoV OR novel CoV OR CoV 2 OR CoV2 OR sarscov2 OR 2019nCoV OR wuhan virus*).mp. OR ((wuhan OR hubei OR huanan) AND (severe acute respiratory OR pneumonia*) AND outbreak*).mp. OR ((wuhan OR hubei OR huanan) AND (coronavir* OR betacoronavir*)).mp.<br><br>Limits: 2019- |

| | |
|---|---|
| Cochrane Library | #1(coronavir* OR "corona virus" OR betacoronavir* OR covid19 OR "covid 19" OR nCoV OR "CoV 2" OR CoV2 OR sarscov2 OR 2019nCoV OR "novel CoV" OR "wuhan virus"):ti,ab,kw OR ((wuhan OR hubei OR huanan) AND ("severe acute respiratory" OR pneumonia*) AND outbreak*):ti,ab,kw<br>#2MeSH descriptor: [Coronavirus] this term only<br>#3MeSH descriptor: [Coronavirus Infections] this term only<br>#4MeSH descriptor: [Betacoronavirus] this term only<br>#5 #1 OR #2 OR #3 OR#4<br>Limits: 2020-<br><br>OR<br><br>#1 ( "novel coronavirus" OR "novel corona virus" OR covid19 OR "covid 19" OR nCoV OR "novel CoV" OR "CoV 2" OR CoV2 OR sarscov2 OR 2019nCoV OR "wuhan virus"):ti,ab,kw OR ((wuhan OR hubei OR huanan) AND ("severe acute respiratory" OR pneumonia*) AND outbreak*):ti,ab,kw OR ((wuhan OR hubei OR huanan) AND (coronavir* OR betacoronavir*)):ti,ab,kw<br>Limits: 2019- |
| Scopus 1960- | TITLE-ABS-KEY ( coronavir* OR "corona virus" OR betacoronavir* OR covid19 OR "covid 19" OR ncov OR "CoV 2" OR cov2 OR sarscov2 OR 2019ncov OR "novel CoV" OR "wuhan virus" ) OR ( TITLE-ABS-KEY ( wuhan OR hubei OR huanan ) AND TITLE-ABS-KEY ( "severe acute respiratory" OR pneumonia* ) AND TITLE-ABS-KEY ( outbreak* ) ) AND ( LIMIT-TO ( PUBYEAR , 2020 ) )<br><br>OR<br><br>TITLE-ABS-KEY ( "novel coronavirus" OR "novel corona virus" OR covid19 OR "covid 19" OR ncov OR "CoV 2" OR cov2 OR sarscov2 OR 2019ncov OR "novel CoV" OR "wuhan virus" ) OR TITLE-ABS-KEY ( ( wuhan OR hubei OR huanan ) AND ( "severe acute respiratory" OR pneumonia* ) AND outbreak* ) OR TITLE-ABS-KEY ( ( wuhan OR hubei OR huanan ) AND ( coronavir* OR betacoronavir* ) ) AND ( LIMIT-TO ( PUBYEAR , 2020 ) OR LIMIT-TO ( PUBYEAR , 2019 ) ) |
| Academic Search Complete (Ebsco) | TI,AB,SU( (coronavir* OR "corona virus" OR betacoronavir* OR covid19 OR "covid 19" OR nCoV OR "CoV 2" OR CoV2 OR sarscov2 OR 2019nCoV OR "novel CoV" OR "wuhan virus") OR ((wuhan OR hubei OR huanan) AND ("severe acute respiratory" OR pneumonia*) AND (outbreak*)) )<br><br>Limits: Dec. 2019-, peer-reviewed |
| Africa Wide Information (Ebsco) | TI,AB,SU( (coronavir* OR "corona virus" OR betacoronavir* OR covid19 OR "covid 19" OR nCoV OR "CoV 2" OR CoV2 OR sarscov2 OR 2019nCoV OR "novel CoV" OR "wuhan virus") OR |

| | |
|---|---|
| | ((wuhan OR hubei OR huanan) AND ("severe acute respiratory" OR pneumonia*) AND (outbreak*)) )<br><br>Limits: 2020-, peer-reviewed<br><br>OR<br><br>TI,AB,SU( ("novel coronavirus" OR "novel corona virus" OR covid19 OR "covid 19" OR nCoV OR "CoV 2" OR CoV2 OR sarscov2 OR 2019nCoV OR "novel CoV" OR "wuhan virus") OR ((wuhan OR hubei OR huanan) AND ("severe acute respiratory" OR pneumonia*) AND outbreak*) OR ((wuhan OR hubei OR huanan) AND (coronavir* OR betacoronavir*)) )<br><br>Limits: 2019-, peer-reviewed |
| CINAHL (Ebsco) | TI,AB,SU( (coronavir* OR "corona virus" OR betacoronavir* OR covid19 OR "covid 19" OR nCoV OR "CoV 2" OR CoV2 OR sarscov2 OR 2019nCoV OR "novel CoV" OR "wuhan virus") OR ((wuhan OR hubei OR huanan) AND ("severe acute respiratory" OR pneumonia*) AND (outbreak*)) ) OR (MH "Coronavirus") OR (MH "Coronavirus Infections")<br><br>Limits: Dec. 2019-, peer-reviewed |
| ProQuest Central (Proquest) 1952- | TI,AB,SU( (coronavir* OR "corona virus" OR betacoronavir* OR covid19 OR "covid 19" OR nCoV OR "CoV 2" OR CoV2 OR sarscov2 OR 2019nCoV OR "novel CoV" OR "wuhan virus") OR ((wuhan OR hubei OR huanan) AND ("severe acute respiratory" OR pneumonia*) AND (outbreak*)) )<br><br>Limits: Dec. 2019-, peer-reviewed |
| PubMed Central | TITLE-ABSTRACT( (coronavirus OR "corona virus" OR coronavirinae OR coronaviridae OR betacoronavirus OR covid19 OR "covid 19" OR nCoV OR "CoV 2" OR CoV2 OR sarscov2 OR 2019nCoV OR "novel CoV" OR "wuhan virus" ) OR ((wuhan OR hubei OR huanan) AND ( "severe acute respiratory" OR pneumonia ) AND (outbreak)) ) OR "COVID-19" [Supplementary Concept] OR "severe acute respiratory syndrome coronavirus 2" [Supplementary Concept]<br><br>Limits: Dec. 2019- |
| Medline (PubMed) | TITLE-ABSTRACT( ( coronavirus OR "corona virus" OR coronavirinae OR coronaviridae OR betacoronavirus OR covid19 OR "covid 19" OR nCoV OR "CoV 2" OR CoV2 OR sarscov2 OR 2019nCoV OR "novel CoV" OR "wuhan virus" ) OR ((wuhan OR hubei OR huanan) AND ( "severe acute respiratory" OR pneumonia ) AND (outbreak)) ) OR "COVID-19" [Supplementary Concept] OR "severe acute respiratory syndrome coronavirus 2" [Supplementary Concept]<br><br>Limits: Dec. 2019- |

| | |
| --- | --- |
| LitCovid (NLM) | https://www.ncbi.nlm.nih.gov/research/coronavirus/ |
| SciFinder (CAS) | References ( coronavir* OR "corona virus" OR betacoronavir* OR covid19 OR covid OR nCoV OR "CoV 2" OR CoV2 OR sarscov2 OR 2019nCoV OR "novel CoV" OR "wuhan virus" )<br><br>Limits: 2020-<br><br>OR<br><br>References ( "novel coronavirus" OR "novel corona virus" OR covid19 OR covid OR nCoV OR "CoV 2" OR CoV2 OR sarscov2 OR 2019nCoV OR "novel CoV" OR "wuhan virus" )<br><br>Limits: 2019- |
| Virtual Health Library (WHO) | Filter VHL created:<br>https://bvsalud.org/vitrinas/post_vitrines/novo_coronavirus/<br>**Database changed on 6/16/2020 and integrated into the larger WHO database**<br>Limited: 2019-<br><br>OR<br><br>TI,AB:( (coronavirus OR "corona virus" OR coronavirinae OR coronaviridae OR betacoronavirus OR covid19 OR "covid 19" OR nCoV OR "CoV 2" OR CoV2 OR sarscov2 OR 2019nCoV OR "novel CoV" OR "wuhan virus" ) OR ((wuhan OR hubei OR huanan) AND ("severe acute respiratory" OR pneumonia) AND (outbreak)) )<br><br>Limits: 2020-<br><br>OR<br><br>TI,AB( "novel coronavirus" OR "novel corona virus" OR covid19 OR "covid 19" OR nCoV OR "CoV 2" OR CoV2 OR sarscov2 OR 2019nCoV OR "novel CoV" OR "wuhan virus") OR ((wuhan OR hubei OR huanan) AND ("severe acute respiratory" OR pneumonia) AND outbreak) OR ((wuhan OR hubei OR huanan) AND (coronavirus OR betacoronavirus))<br><br>Limits: 2019- |
| WHO Novel Coronavirus page | Download of their global research on COVID 19 database:<br>https://www.who.int/emergencies/diseases/novel-coronavirus-2019/global-research-on-novel-coronavirus-2019-ncov |

| | |
|---|---|
| | OR<br><br>Hand search |
| CDC Novel Coronavirus page | https://www.cdc.gov/coronavirus/2019-ncov/publications.html<br><br>OR<br><br>Hand search |
| EuroSurveillance | https://www.eurosurveillance.org/content/2019-ncov?pageSize=100&page=1 |
| China CDC MMWR | Hand search |
| Homeland Security Digital Library | Title or Summary: Coronavirus OR "Corona virus" OR Betacoronavirus OR Coronaviridae OR coronavirinae OR Covid OR Covid19 OR nCoV OR CoV OR CoV2 OR Wuhan<br><br>Limits: 2019- |
| ClinicalTrials | Condition, disease, other term: Coronavirus OR "Corona virus" OR Betacoronavirus OR Coronaviridae OR coronavirinae OR Covid OR Covid19 OR nCoV OR CoV OR CoV2 OR Wuhan<br><br>Limits: 2019- |
| bioRxiv medRxiv chemRxiv (preprints) | Condition, disease, other term: Coronavirus OR "Corona virus" OR Betacoronavirus OR Coronaviridae OR coronavirinae OR Covid OR Covid19 OR nCoV OR CoV OR CoV2 OR Wuhan<br><br>Limits: 2019- |
| SSRN (preprints) | Condition, disease, other term: Coronavirus OR "Corona virus" OR Betacoronavirus OR Coronaviridae OR coronavirinae OR Covid OR Covid19 OR nCoV OR CoV OR CoV2 OR Wuhan<br><br>Limits: 2019- |
