## Supplementary Material Online 2 for "Clinical Trials in COVID-19 Management & Prevention: A Meta-epidemiological Study examining methodological quality"

**Supplemental Material Online 2.** Modified Cochrane Risk of Bias Tool assessment criteria.

| Bias from randomization process<br>[Considerations: Random sequence generation, allocation concealment] | |
|---|---|
| Definitely low risk of bias | Trials that assign participants to alternative interventions using a randomly generated sequence and maintain allocation concealment. *An explicit description of random sequence generation is not necessary for a rating of low risk of bias.* |
| Probably low risk of bias | Trials in which healthcare providers were blind to the intervention but which provide no information on allocation concealment and in which there are no major baseline imbalances. *An explicit description of random sequence generation is not necessary for a rating of probably low risk of bias.* |
| Probably high risk of bias | Trials in which healthcare providers were not blind to the intervention and which provide no information on allocation concealment **OR** trials in which there are substantial baseline differences between trial arms that suggest a problem with the randomization process but there are no other limitations related to randomization. |
| Definitely high risk of bias | Trials in which allocation is by judgment of the clinician, by preference of the participant, by availability of the intervention, based on the results of a laboratory test, or other non-random rules (e.g., birthdate, etc.) **OR** trials in which investigators enrolling participants could possibly foresee the arm to which each subsequent patient would be randomized, such as allocation using an open allocation schedule (e.g. a list of random numbers), assignment envelopes used without appropriate safeguards (e.g. use of unsealed, non-opaque or not sequentially numbered envelopes), alternation between arms, case record number, or any other explicitly unconcealed procedure, rate as high risk. |
| **Bias due to deviations from intended intervention**<br>[Considerations: Blinding of clinicians & participants, imbalances in co-interventions or behaviours] | |
| Definitely low risk of bias | Therapy trials in which healthcare providers are blind to the intervention administered and in which there are no significant differences in administered co-interventions **OR** therapy trials that are described as double or triple blind. |
| Probably low risk of bias | |
| Probably high risk of bias | Therapy trials in which healthcare providers are not blind to the intervention administered **OR** therapy trials in which healthcare providers are blind to the intervention administered but there are significant differences in administered co-interventions that suggests that blinding may have been compromised **OR** therapy trials in which healthcare providers are described as being blind to the intervention but allocation concealment was inadequate. d |
| Definitely high risk of bias | Therapy trials in which healthcare providers are not blind to the intervention and in which there are significant differences in administered co-interventions. |
| **Bias due to missing data**<br>[Considerations: Missing outcome measures, loss to follow-up] | |
| Definitely low risk of bias | Trials in which missing outcome data (including outcome data that has been imputed) < 10%. |
| Probably low risk of bias | Trials in which missing outcome data (including outcome data that has been imputed) is between 10% to 15% and missing outcome data is unlikely to be related to the true outcome and there is no imbalance in numbers of or reasons for missing data across intervention groups. |
| Probably high risk of bias | Trials in which missing outcome data (including outcome data that has been imputed) is between 10% to 15% and missing outcome data is likely to be related to the true outcome or there are imbalances in numbers of or reasons for missing data across intervention groups. |

| | |
|---|---|
| Definitely high risk of bias | Trials in which missing outcome data (including outcome data that has been imputed) > 15%. |
| **Bias due to measurement of the outcome**<br>[Considerations: Blinding of outcome adjudicators, objectivity of outcome] | |
| Definitely low risk of bias | Trials in which (1) patients are blind to the intervention and in which outcomes are patient-reported, or (2) outcomes are measured by a third-party (investigator or clinician) and in which the third-party is blind to the intervention, or (3) the outcomes are objective, or (4) described as double or triple blind. |
| Probably low risk of bias | |
| Probably high risk of bias | |
| Definitely high risk of bias | Trials in which patients are not blind and in which outcomes are patient-reported (e.g., time to symptom resolution) or trials in which outcome adjudicators are not blind and the outcomes are not objective. |
| **Bias in selection of the reported results**<br>[Considerations: Selective reporting of outcome measures & timepoints] | |
| Definitely low risk of bias | Results for outcomes that were analyzed and reported according to a pre-specified statistical analysis plan or protocol (including the timepoint for the measurement of the outcome). |
| Probably low risk of bias | Results for outcomes that were analyzed and reported but that were not prespecified in a statistical analysis plan or protocol but the timepoint at which results are reported is consistent with the timepoint for other outcomes in the trial report or there is little reason to believe the outcome was selectively reported. |
| Probably high risk of bias | Results for outcomes that were analyzed and reported but that were not prespecified in a statistical analysis plan or protocol but the timepoint at which results are reported is not consistent with the timepoint for other outcomes in the trial report or there are other reasons to believe that the outcome is selectively reported. |
| Definitely high risk of bias | Results for outcomes that were analyzed and reported for which there are inconsistencies with the statistical analysis plan or protocol. These inconsistencies may include outcome measures of interest or the timepoints for the measurement of outcomes. |
| **Other bias**<br>[Considerations: Competing risks due to early termination for continuous outcomes only] | |
| Definitely low risk of bias | Results are very unlikely to have been affected by competing risk due to death. |
| Probably low risk of bias | Results are unlikely to have been affected by competing risk due to death. |
| Probably high risk of bias | Results are likely to have been affected by competing risk due to death. |
| Definitely high risk of bias | Results are very likely to have been affected by competing risk due to death. |
